# Country distancing reveals the effectiveness of travel restrictions during COVID-19

**DOI:** 10.1101/2020.07.24.20160994

**Authors:** Lu Zhong, Mamadou Diagne, Weiping Wang, Jianxi Gao

## Abstract

Travel restrictions are the current central strategy to globally stop the transmission of the novel coronavirus disease (COVID-19). Despite remarkably successful approaches in predicting the spatiotemporal patterns of the ongoing pandemic, we lack an intrinsic understanding of the travel restriction’s effectiveness. We fill this gap by developing a surprisingly simple measure, *country distancing*, that is analogical to the *effective resistance* in series and parallel circuits and captures the propagation backbone tree from the outbreak locations globally. This approach enables us to map the effectiveness of travel restrictions to arrival time delay (ATD) or infected case reduction (ICR) systematically. Our method estimates that 50.8% of travel restrictions as of Apr-4 are ineffective, resulting in zero ATD or ICR worldwide. Instead, by imposing Hubei’s lockdown on Jan-23 and national lockdown on Feb-8, mainland China alone leads to 11.66 [95% credible interval (CI), 9.71 to 13.92] days of ATD per geographic area and 1,012,233 (95% CI, 208,210 −4,959,094) ICR in total as of Apr-4. Our result unveils the trade-off between the *country distancing* increase and economic loss, offering practical guidance for strategic actsion about when and where to implement travel restrictions, tailed to the real-time national context.

---

The COVID-19 with 8,860,331 confirmed cases and 465,740 deaths worldwide, as of June 22, 2020, was first reported to the WHO (World Health Organization) on December 31, 2019 *(1, 2)*. Today’s high population density *(3)* and high volume, speed, and nonlocality of human mobility provide perfect conditions for epidemic spreading *(4, 5)*, and simultaneously raise the challenges for non-pharmaceutical intervention strategies on the time scale at the pace modern diseases can spread *(6, 7)*. Specifically, through the global mobility network (GMN), mainland China introduces 288 infected cases to other geographic areas from January 3 to February 13, 2020 *(8, 9)*. As COVID-19 is declared a pandemic on March 11, 2020, more than half of the world was infected, and geographic areas that are continuously exposed to massive airline transits from different infected countries are currently in high importation risk *(7)*. For example, the virus in the US is mainly imported from European countries, including France, Austria, and the Netherlands *(10, 11)*, which is experiencing an upper infection with 2,241,178 confirmed cases and 119,453 deaths till June 22, 2020.

Although the practice of quarantine and social distancing protocols can drastically reduce its propagation locally *(12)*, the global pandemic patterns of COVID-19 are shaped by the GMN, which determines when and where the disease arrives globally *(13)*. Consequently, the straightforward way for lowering the international importation of COVID-19 is to impose radical travel restrictions (i.e., entry ban, global travel ban, and lockdown) *(14,15)* to shrink the entry of airline passengers. As of April 4, 2020, 187 geographic areas imposed the entry ban, 87 geographic areas imposed the global travel ban, and 70 geographic areas imposed the lockdown to prevent their citizens and tourists from traveling overseas *(16, 17)*. However, researchers demonstrated that these travel restrictions are only effective at the beginning of an outbreak *(18)*. Moreover, they would interrupt the healthcare aid and technical support, disrupt businesses, and cause extensive and profound social and economic damage *(19,20)*. Therefore, it is crucial to assess and impose effective travel restrictions to avoid uncoordinated government responses to COVID-19, which may lead to a substantial unnecessary cost.

Measuring the travel restrictions’ effectiveness often relies on the specific epidemic models *(21,22)*, which require accurate estimation of the disease’s epidemiological parameters, such as the basic reproductive number (R0). In spite of that, the parameter estimations are often not reliable due to the daily changing under-reported cases and various errors due to insufficient diagnosis tests *(6, 23, 24)*. Furthermore, these models are hard to calibrate due to incomplete information (i.e., partial network topology *(25)* or unknown dynamics *(14, 26)*), and it is unclear how much details are required to achieve a certain level of predictive accuracy. Human mobility plays a crucial role in understanding the hidden spatiotemporal spreading patterns and enables us to predict the arrival time *(13)* and estimate the number of infected cases *(27)* without knowing the epidemiological parameters. It is remarkable that despite the complex topology of the mobility network, a dominant trajectory defined as the effective distance *(13)* can always be identified from the outbreak location (OL) to the target geographic area by discarding other redundant connections. This method reliably predicts the arrival time and epidemic wavefront without knowing the epidemiological parameters, which has already been demonstrated in both the pandemic H1N1 and the global 2003 SARS epidemic. On the other hand, the aggregate mobility outflow from the OL has also been a vital predictor for the cumulative number of infections in the destination location *(27)*, validated by the Wuhan’s outflow to each prefecture in mainland China. Despite advances of both approaches and their follow-up methods *(7, 28)*, they are more suitable for the early stage of the pandemic of COVID-19 than the late stage when multiple OLs arise, increasing the level of complexity that promotes the needs of new mathematical tools.

## Global Disease Transmission Law and Country Distancing

We test the international spreading of the COVID-19 on the global mobility network (GMN) *G* = *(N, E, F*) (See Methods, Tab. S1 and Tab. S2) provided by the Official Aviation Guide *(29)*, where *N* denotes geographic area set, *E* denotes the airline link set, and *F*_*mn*_ *(F*_*mn*_ *∈ F*) represents a weighted value describing the airline passenger influx from area *n* to the area *m*. To eliminate the effect of geographic areas’ sizes, we define 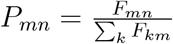 as the fraction of individuals that leave *n* and go to *m*. For a complex GMN with a single OL, the diseases may propagate to the destination through different paths. But the arrival time only depends on the shortest path whose length is defined as the *effective distance (13)*, since arrival time equals the *effective distance* divided by the effective spreading velocity v_eff_. The *effective distance* from *n* to a connected node *m* is defined as *d*_*m*|*n*_ = 1 − log *P*_*mn*_, derived from the likelihood of transmitting the disease from *n* to *m*, that is 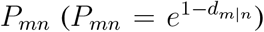. For an arbitrary node that can be reached by *n* through a path *τ* = {*n*,…, *i, j*,…, *m*}, the effective distance is the sum of effective lengths along the links of the shortest path, 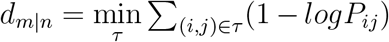. Thus, we obtain the series law for global disease transmission:

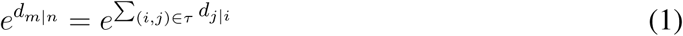

We call this series law because it is analogical to the *effective resistance* in series circuits defined as *R* = ∑*i R*_*i*_, where *R*_*i*_ is the resistance that is connected along a chain. Take the series connection of resistance as an example in Fig. 1A. The *effective distance* from *n* to *m* in GMN satisfies 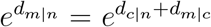, that is, *d*_*m*|*n*_ = *d*_*c*|*n*_ + *d*_*m*|*c*_, where *c* is on the shortest path between *m* and *n*, which is analogical to *R* = *R*_*n*_ + *R*_*c*_.

**Figure 1:**
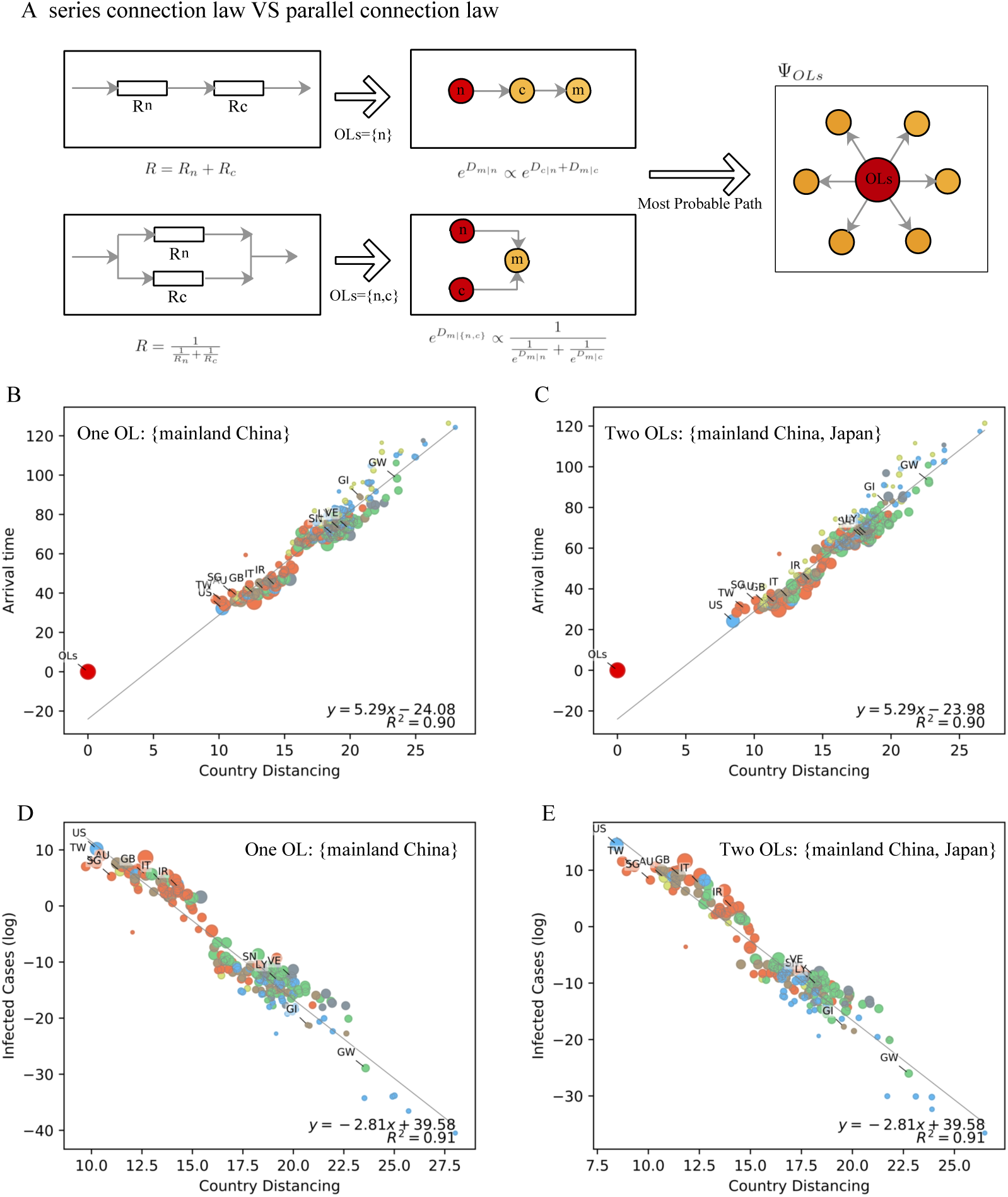
Understanding the *country distancing* and its correlation with arrival times and infected cases in the presence of one and multiple outbreak locations. (A) Description of series and parallel laws. (B)(D) Correlation between *country distancing* and simulated arrival times (B) and correlation between *country distancing* and simulated infected cases in log transformation (D) when the defined OL set *(N*_*I*_) is {mainland China}. The arrival times and infected cases are simulated with the meta-population susceptible-infected-recovered (SIR) model *(31)* with the given epidemiological parameters of COVID-19 *(32)* (see supplementary text). (C)(E) Correlation between *country distancing* and arrival times (C) and correlation between *country distancing* and infected cases (E) when the defined OL set *(N*_*I*_) is {mainland China, Japan}. Geographic areas are represented by circles. Those in the same continent are filled in the same color.

Next, we develop the parallel law for global disease transmission when multiple OLs present. For example, the disease propagates from OLs, *n* and *c*, to the destination geographic area, *m*, with *effective distances d*_*m*|*n*_, and *d*_*m*|*c*_ respectively, as shown in Fig. 1A. Therefore, the likelihoods of transmitting disease from *n* and *c* are 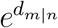 and 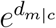 respectively. Thus, the overall likelihood of transmitting from both OLs satisfies 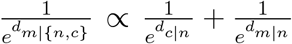 (See methods and supplementary text). This process is similar to the *effective resistance* in parallel circuits that 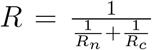. For the general case, we derive the parallel law for global disease transmission and formulate the *country distancing* of area *m* from the set of OLs *N*_*I*_ as (see supplementary text)

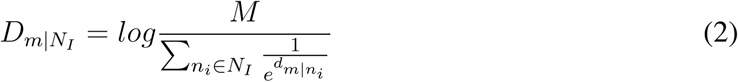

where *N*_*I*_ = {*i*|*∀i ∈ N* &*I*_*i*_ > *I*_c_} is the outbreak location set at time *t*. Here, we set *I*_c_ = 100, as it is observed that the confirmed cases of COVID-19 raise exponentially after the 100th case is confirmed *(30)*. *I*_*i*_ is the accumulative confirmed infected cases at area *i* and |*N*_*I*_| ≤ *M*. See supplementary text [Eqs. (S3)-(S12)] for the details about Eq. (2). Note that a large set of OLs *N*_*I*_ may lead to a small distance 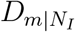, because the more OLs, the higher probability to arrive at geographic area, *m*. Our series and parallel law for global disease transmission enable us to map any complex GMN to a star-like network, capturing the propagation backbone tree from the outbreak locations globally, such as the shortest path tree Ψ_*OLs*_ depicted in Fig1. A.

Two fundamental properties – the arrival time *(T*_*m*_) and the infected cases *(I*_*m*_) in arbitrary geographic area *m* – describe the major spread patterns of the pandemic of COVID-19. Increasing findings show the strong correlation between human mobility and arrival times *(7, 13, 28)* for the propagation of H1N1 and SARS, and the strong correlation between mobility flows from Wuhan, mainland China and infected cases in other cities in mainland China *(27)* for the propagation of COVID-19. Therefore, human mobility determines both fundamental properties of the spreading patterns, although they are established in the presence of a single OL. Motivated by these independent but correlated researches together with our series and parallel law for global disease transmission, we surprisingly find that *country distancing* generates linear relationships with the arrival times 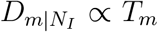, and the logarithm of the infected cases 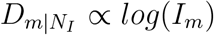, as shown in Fig. 1B-E and Fig. S1:

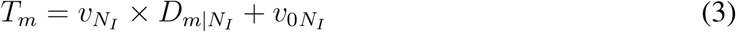

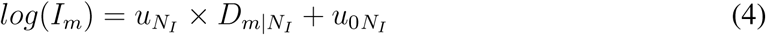

where 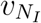and 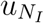 are the slopes, respectively, representing the rates of change of arrival times/infected cases relative to *country distancing*.

We simulate the spread of COVID-19 by adopting the meta-population susceptible-infected-recovered (SIR) model *(31)* with the given epidemiological parameters of COVID-19 *(32)*. As shown in Fig. 1B and C, when mainland China is the only OL, i.e., *N*_*I*_ = {mainland China}, we observe a strong correlation between *country distancing* and arrival times with *R*^2^ = 0.90 and 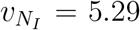 (95% CI, 4.52 to 6.01), indicating a 5.29 days delay of arrival time at area *m* for an increase of 1 in 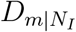 due to travel restrictions. For the correlation between *country distancing* and infected cases, *R*^2^ = 0.91 and 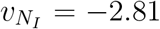 (95% CI, −3.23 to −2.38). It means that 94% (1 − *e*^2.81^) of infected cases are reduced at area *m* for an increase of 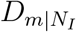 due to travel restrictions. Furthermore, when *N*_*I*_ = {mainland China, Japan} as shown in Fig. 1D and E, *R*^2^ = 0.90 and slope 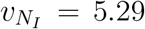 (95% CI, 4.53 to 6.01) for the correlation between *country distancing* and arrival times and *R*^2^ = 0.91 and slope 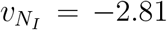 (95% CI, −3.26 to −2.40) for the correlation between *country distancing* and infected cases. For the evolving OL set according to the COVID-19 time-series data provided by Johns Hopkins University and Ding Xiang Doctor Website (see Methods), we also systematically test the sensitivity of the analysis (Fig. S3). By using the *country distancing*, we could give the generic spreading speed 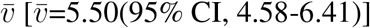, and generic spreading rate, *ū*, [*ū*=-2.95 (95% CI,-3.51 to −2.38)], for the different set of OLs [see (Fig. S3)]. Consequently, we can translate the changes of *country distancing* to the arrival time delay (ATD) and infected case reduction (ICR) with Eqs. (15) and (16).

### Status Quo of Travel Restrictions

Available Dataset reveals that 249 geographic areas implement a total of 663 travel restrictions as of Apr-4. A total number of 476 entry bans imposed by 184 countries (73.8%) from Jan-21 to March-17. These areas which imposed entry bans deny access to non-citizens who have been at some specific geographical regions such as mainland China, South Korea, Japan, Iran, Schengen Area. At the early stage from Jan-21 to Feb-6, 49 areas (19.6%) imposed entry bans to distance themselves from the initially infected area, namely, mainland China. As the COVID-19 continued spreading outside of mainland China, most areas extended their entry bans to South Korea (42%), Japan (64%), Iran (4%), and Schengen Area (5.6%) from Feb-8 to Mar-13. Knowing that mainland China imposed lockdown at Hubei Province (Jan-23) and national lockdown (Feb-8) to distance itself from the world, few areas imposed an entry ban to mainland China since Feb-8. From Mar-11, when about half of the countries were infected, worldwide, entry bans were not sufficient to lower the risk of coronavirus importation from the infected regions. Consequently, 87 areas (34.9%) imposed the global travel ban to prevent oversea travels from entering their areas except for their residents, and 65 countries (26.1%) imposed the partial lockdowns and 12 (4.8%) countries imposed full/national lockdowns to prevent people entering and exiting their countries (see Fig. 2A-B and Tab. S3).

**Figure 2:**
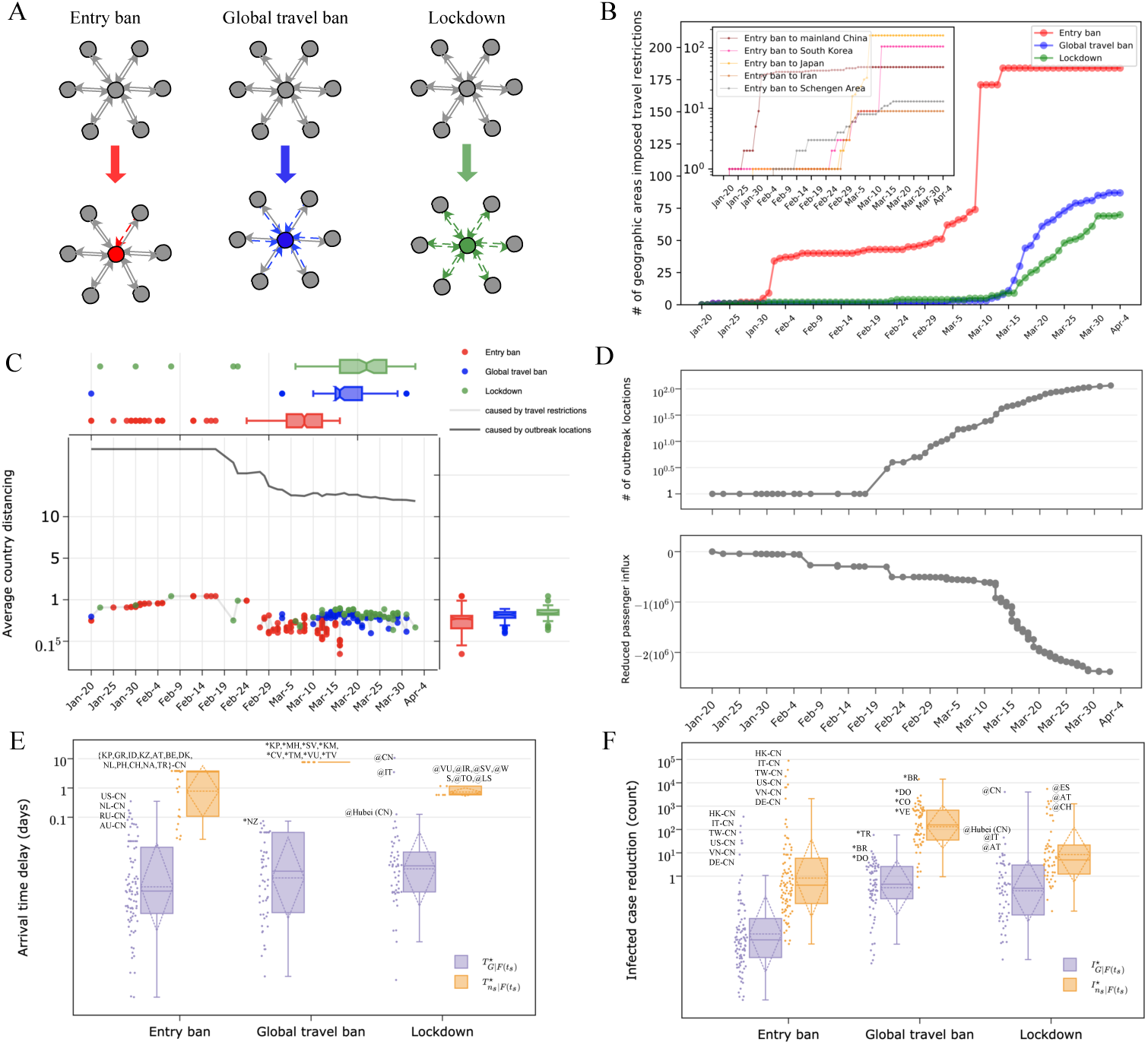
*Country distancing*, outbreak locations (OLs), and three types of travel restrictions (i.e., entry ban, global travel ban, lockdown) from Jan-20 to Apr-4. (A) Illustration of how the entry ban, global travel ban, and lockdown reduce airline passenger influx. When the colored areas (in red, blue, and green) enforce the entry ban, the global travel ban, or the lockdown, the passenger influx on corresponding links are reduced (solid links change to dotted links). (B) Number of geographic areas that imposed travel restrictions as of Apr-4. The embedding subplot presents the number of geographic areas that imposed entry bans to the main five areas. (C) Average *country distancing* 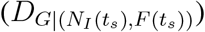. Here, 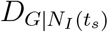 (black line) is the average *country distancing* resulting from OLs, and 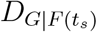 (colored dots) is the average *country distancing* resulting from travel restrictions. (D) Growth of outbreak locations |*N*_*I*_*(t*_*s*_)|, and the decline of passenger influx *F (t*_*s*_) caused by travel restrictions *s* ∈ 𝕊. (E) Arrival time delay (ATD) caused by travel restrictions. (F) Infected case reduction (ICR) caused by travel restrictions. In (E)(F), ATD and ICR are visualized for two groups, that is, the average of ATD 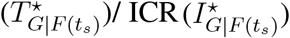 for the world, and the ATD 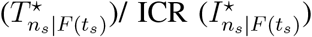 of geographic area *n*_*s*_ imposed *s*^*th*^ travel restriction. Among the marks, for example, “US-CN” represents the entry ban imposed by US to CN (mainland China); “*KP” represents the global travel ban imposed by North Korea; “@CN” represents the lockdown imposed by mainland China. For clear visualization, geographic areas are presented by two-letter code.

### Effectiveness of Travel Restrictions

The existing travel restrictions that are imposed to curb the spread of COVID-19 (see Fig.2 A-B and Tab. S3) are presented by a quadruple {𝕊, 𝕋, ℕ, 𝔼}. The *s*^*th*^ *(s* ∈ 𝕊) travel restriction, which is imposed by the geographic area *n*_*s*_ *∈* ℕ at time *t*_*s*_, reduces the passenger influx of airline links in *E*_*s*_ ∈ 𝔼 with different strengths (See Methods for details). As illustrated in Fig.2 A, the entry ban only reduces the passenger influx to banned areas from area *n*_*s*_; the global travel ban reduces the passenger influx from all neighbor areas to enter the area *n*_*s*_; the lockdown reduces the passenger influx entering/leaving the area *n*_*s*_.

Notice that the *country distancing* decreases with the number of OLs and increases with the reduction of passenger influx induced by travel restrictions. Therefore, we consider the number of OLs and travel restriction as two separate factors that influence the changes of *country distancing* over time, 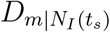 and 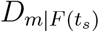, where 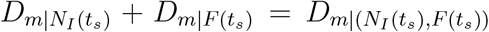 [see Eqs. (8)-(14)]. Fig.2C shows the influence of OLs and travel restrictions on *country distancing* from Jan-20 to Apr-4. From the early stage until Feb-22, mainland China is the only OL. After Feb-22, an increase of the number of OLs occurs, |*N*_*I*_*(t*_*s*_)| > 1, and the average *country distancing* 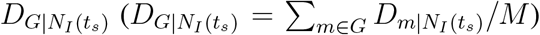 drops from 18.13 to 16.5. As of Apr-4, |*N*_*I*_*(t*_*s*_)| increases to 119 areas, leading to a consecutive decrease in *D*_*G*|*N*_*I (ts)* to 11.8. Concurrently, the travel restrictions imposed before Feb-22 reduce a relatively small amount of passenger influx given by *n,m(F*_*nm*_*(t*_0_) − *F*_*nm*_*(t*_*s*_)) as shown in Fig. 2D, and the average *country distancing* 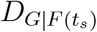 resulting from travel restrictions grows to 2.71 as shown in Fig. 2C. After Feb-22, when |*N*_*I*_*(t*_*s*_)| = 5 at Feb-27, 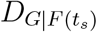 is smaller than 0.1, enabling to conclude that all travel restrictions after Feb-27 are not really effective. This fact also indicates that the *country distancing* is further shortened to the number of OLs |*N*_*I*_*(t*_*s*_)|. Our analysis confirms that the presence of multiple OLs magnifies areas’ risk of coronavirus importation. The risk of importation can only be mitigated by rising substantially the effort of travel restrictions.

Through all the travel restrictions as of Apr-4, the arrival time delay reaches 16.69 days (95% CI, 13.90 to 19.45) on average, and the infected case reduction reaches 1,257,257 (95% CI, 281,475 to 6,051,856) in total, worldwide. We find that 337 (50.8%) of travel restrictions are ineffective, leading to zero average increase in *country distancing*, as shown in Tab. S4. The travel restrictions that lead to < 0.1 of 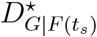 are Hubei province’s lockdown on Jan-23, mainland China’s national lockdown on Feb-8, and Italy’s national lockdown on Feb-23. The rest of 326 travel restrictions lead to nonzero increase in *country distancing* (see Fig. S5 and Fig. S6), and their corresponding ATD and ICR are shown in Fig. 2E-F. Mainland China’s lockdown on Feb-8 results in 10.13 days of average ATD and 3871.22 of average ICR, separately accounting for 60% of total ATD and 76.9% of total ICR. Italy’s lockdown on Feb-23 results in 3.46 of average ATD, accounting for 20.7% of total average ATD. A few entry bans from the {United States (US), Netherlands (NL), Russia (RU), Australia (AU)} to mainland China (CN) results in > 0.1 of average ATD, and no global travel bans result in > 0.1 of average ATD. Similarly, for the ICR, entry bans from {Hong Kong (HK), Italy (IT), Taiwan (TW)} to mainland China results in > 100 of average ICR, and no global travel bans result in > 100 of average ICR. Usually, the areas *n*_*s*_, which imposed travel restrictions have higher ATD and ICR for itself than for other areas. For example, North Korea (KP)’s entry ban to mainland China and North Korea ‘s global travel ban respectively increase 3.81 and 7.62 ATD for North Korea. Hong Kong (HK), Italy (IT), and Taiwan (TW) has most ICR, e.g., 88,346, 35,713, 18,911, for their own areas through entry bans.

Next, we explore what type of travel restrictions are effective and visualize the shortest path tree Ψ_*OLs*_ of three examples of travel restrictions, such as the US’s entry ban to mainland China, North Korea’s global travel ban, and mainland China’s lockdown as shown in Fig. 3A-C. By implementing an entry ban to mainland China on Jan-31, the US increases its own *country distancing* by 0.69 and increases a total of 14.35 *country distancing* for the 26 areas, who are descendants of the US in Ψ_*OLs*_. Explicitly, an average of 3.49 ATD and a total of 0 ICR are for the 26 descendant areas. On the other side, by imposing a global travel ban on Jan-21, North Korea’s *country distancing* rises by 1.38 while induces no *country distancing* change for other countries, because North Korea is a leaf node. Mainland China’s lockdown results in no self *country distancing* change but increases a total of 2109.16 of *country distancing* for all other 227 areas. As all other areas are descendants of mainland China, an OL, an average of 10.13 of ATD, and an average of 3817 ICR occur. Similarly, Italy, another OL, leads to no change in itself *country distancing* but increases a total of 722 *country distancing* for other 226 areas by imposing lockdown on Feb-23. Explicitly, an average of 3.46 ATD and an average of 43.23 occur for other 226 areas. The previous statistics reveal that OLs have the most significant influence when distancing the world because they are the sources of the shortest path tree Ψ_*OLs*_. The effective travel restrictions share the same feature as restricting passengers from outbreak locations entering to other areas. The more passengers from outbreak locations are prevented from entering into more areas, the more effective the travel restriction is. In other words, among the travel restrictions preventing passengers from outbreak locations from exiting, the travel restrictions that are imposed by the areas who have more descendant areas in Ψ_*OLs*_ are more effective.

**Figure 3:**
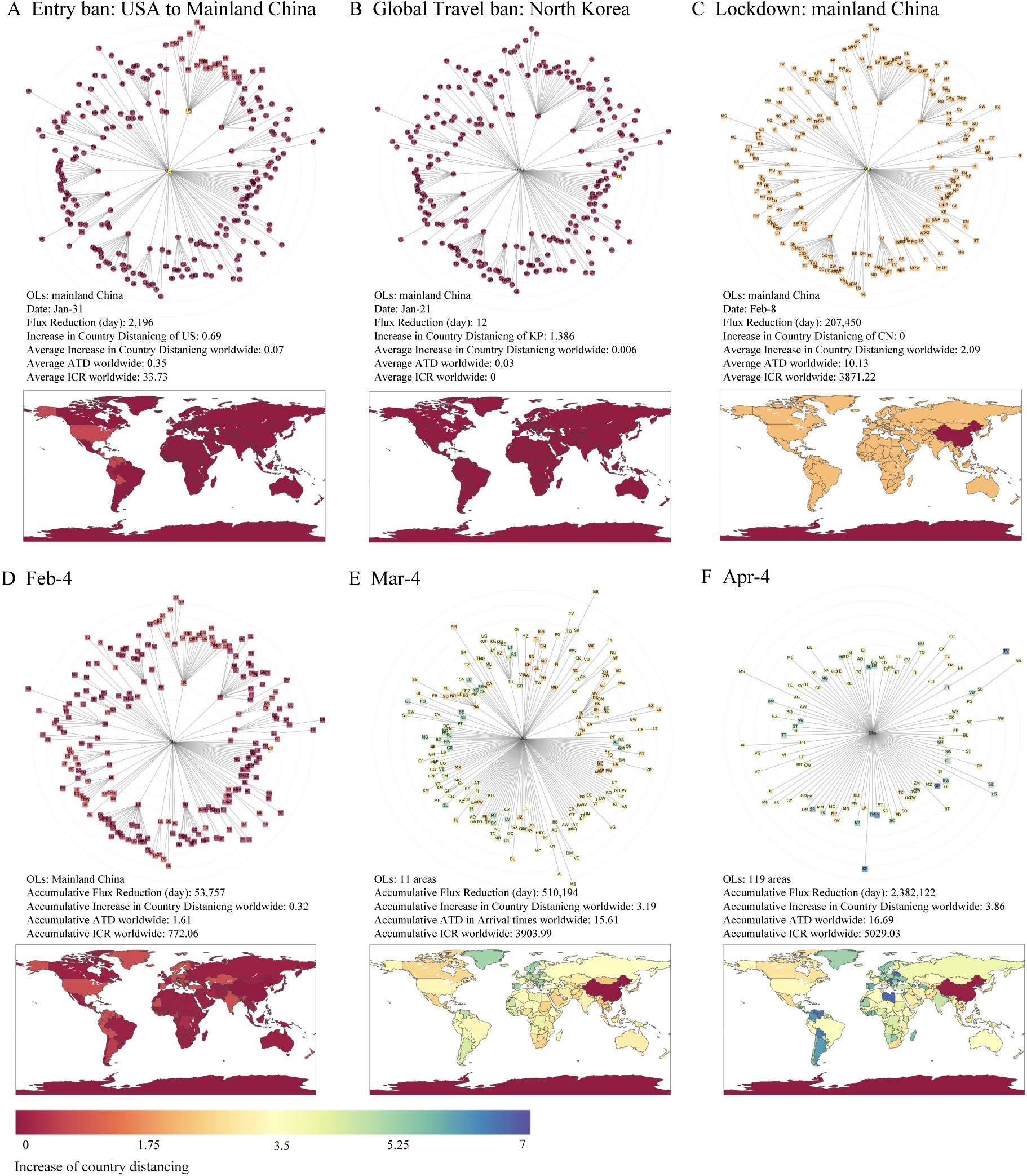
Visualizations of shortest-path tree Ψ_*OLs*_ for the examples of travel restrictions and snapshots of geographic areas with an increase in *country distancing* till Feb-4, Mar-4, and Apr-4 (DEF). (A) Entry ban imposed by the US to mainland China on Jan-31. (B) Global travel ban imposed by North Korea on Jan-21. (C) National lockdown imposed by mainland China on Feb-8. (D)(E)(F) Geographic areas with an increase in *country distancing* as of Feb-4 (D), Mar-4 (E), and Apr-4 (F). Notably, OLs represent the set node of outbreak locations. The radius differs in each shortest path tree.

### Contribution and Benefit via Travel Restrictions

In Fig3.D-F, we show that areas with an increase in *country distancing* till Feb-4, Mar-4, and Apr-4 in the shortest-path tree. Correspondingly, the areas’ ATD 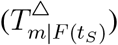 and ICR 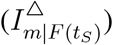 till Feb-4, Mar-4, and Apr-4 are shown in Fig. S7. As of Feb-4, areas and their branch areas increase their *country distancing* mainly by imposing entry bans to mainland China, leading to 0.32 of the average *country distancing* increase. As of Mar-4, the average *country distancing* increase is 3.19 worldwide, mainly due to mainland China’s lockdown and Italy’s lockdown. As of Apr-4, only an additional increase of 0.69 in country distancing occur, proving the inefficiency of travel restrictions in March. We also rank the most beneficial areas due to the increases in *country distancing*, ATD, and ICR, respectively (Tab. S6). Note that Tuvalu (TV), North Korea (KP), and Turkmenistan (TM), which are not infected until Apr-4, are the top three areas with most ATD, i.e., 36.65 days, 35.3 days, and 31.62 days, respectively. Hong Kong (HK), South Korea (KR), and Italy (IT), which are infected at the early stage, are the top three areas with most ICR, i.e., 394,976 cases, 218,800 cases, and 160,766 cases, individually. These findings refute the unconscious statement considering that travel restrictions are only useful for the areas that are not infected. Our analysis suggests that wise travel restrictions imposed at the early stage, could delay the arrival times for uninfected areas and reduce the infected cases for infected areas by continuously lowering the importation risk from OLs.

Since geographic areas implemented different travel restrictions, we integrate the increase in *country distancing* for travel restrictions imposed by same areas *n*_*s*_. We also integrate the ATD and ICR at area *m* due to the travel restrictions imposed by the same areas *n*_*s*_. The ATD and ICR of areas in the same continents brought by area *n*_*s*_’s travel restrictions are shown in Fig. 4. Mainland China has the most significant contributions to the world, accounting for 69% of the increase in *country distancing*, 63.7% of ATD, and 80.5% of ICR. Mainland China’s contribution of ATD is relatively uniformly distributed throughout the six continents, but its contribution of ICR mainly to Asia. Italy gives rise to 23.6% of *country distancing* increase, 18.5% of ATD, and 3.7% of ICR. Italy’s contribution of ATD is to Africa, North Korea, South Korea, and Europe, and its contribution of ICR is mainly to Europe. Hong Kong brings in 0.02% of the increase in *country distancing*, 0.3% of ATD, but an excellent 7.0% of ICR, mainly for Asia. We show the other geographic areas’ contributions in Fig. S8 and Tab. S7. Africa has the longest ATD due to the relatively fewer infected countries in the continent when some significant travel restrictions are implemented, followed by South America, North Korea, Oceania, Europe, and Asia (see Fig. S9). The Asian continent has benefited most from ICR because Asian areas implemented travel restrictions at the early stage to protect themselves, followed by Europe, North America, South America, Oceania, and Africa (see Fig. S9).

**Figure 4:**
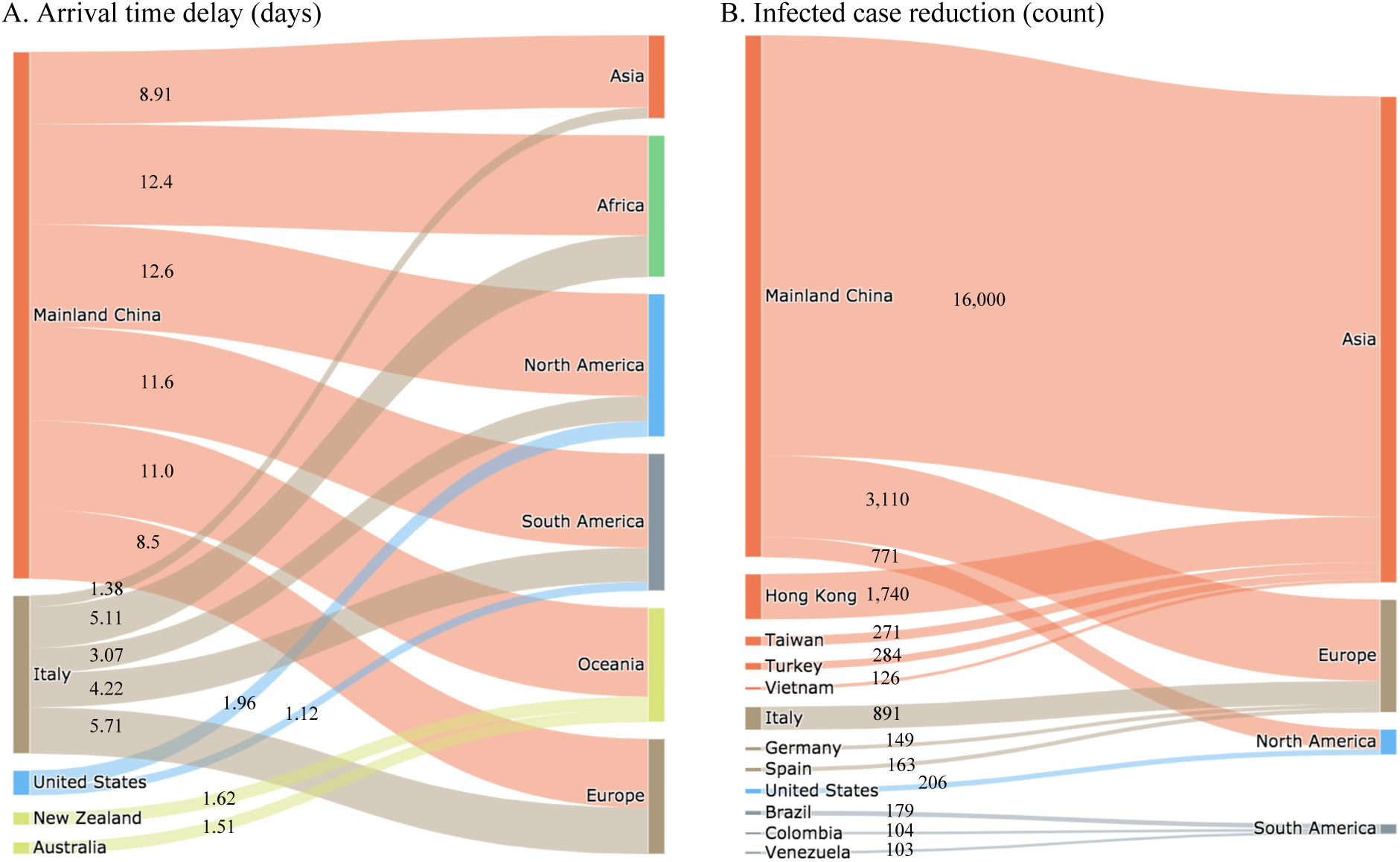
Geographic areas’ contribution in arrival time delay (ATD) and infected case reduction (ICR) to different continents till Apr-4. (A) Arrival time delay (ATD). (B) Infected case reduction (ICR). The flow from the left to the right reflects the ATD/ICR 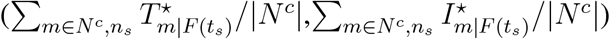 of the right continents for travel restrictions imposed by the left areas *(n*_*s*_). The notation *N*^*c*^ denotes the set of geographic areas in the six continents and *c ∈* {Asia, Africa, North America, South America, Oceania, Europe}. For clear visualization, only the flows with ATD > 1 and ICR > 100 are presented.

### Recommendations for Effective and Economic Travel Restrictions

After a comprehensive analysis of existing travel restrictions, we find that most of them are inefficient for two reasons: (1) the travel restrictions are imposed by geographic areas in an uncoordinated way out of self-interest, failing to contribute to global good; (2) the sole travel restriction is not enacted in optimal time and optimal locations for the most significant self-interest. Lacking the consideration of imposing the coordinated travel restrictions for the global good and strategical plan of when and where to impose each travel restriction, though existing travel restrictions contribute to some increase in ATD and ICR, they are far from slowing the spread of COVID-19. Severely, these travel restrictions created a huge unnecessary loss of passenger influx, ultimately damaging the global economy and social instability *(20, 33)*.

To address the ineffectiveness and inefficiency of the existing travel restrictions, we formulate it as a bi-objective optimization problem: maximizing the travel restrictions’ increase in *country distancing* and minimizing the loss of airline passenger influx in GMN [see Eq. (20) in Methods]. For each travel restriction *s* ∈ 𝕊, which reduces passenger influx of airline links in set *E*_*s*_, we find its optimal solution *s*^*t*^, which reduces passenger influx of each airline link in set 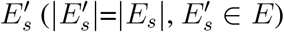 with the strength *α*, like a set of entry bans. We use the Non-dominated Sorting Genetic Algorithm (NSGA-II) to obtain non-dominated solutions for each travel restriction and present the approximate optimal solution, which has the largest 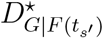 in Fig. 5A-C. Our results show that the optimized travel restrictions significantly outperform the existing travel restrictions in the remaining passenger influx, the average arrival time delay (ATD), and the average infected case reduction (ICR). Form Jan-20 to Feb-20, when mainland China is the single outbreak location (shaded region in Fig. 5A-C), existing travel restrictions result in 12.03 days of average ATD and 4718 cases of average ICR by reducing 295,957 passenger influx (95.9% of passenger influx remains). The optimal travel restrictions reach the ATD of average 17.86 days (vs. 12.03 days, 48% larger) and ICR of average 593,501 (vs. 4,718, 123 times larger) cases for the world. Besides, the optimal travel restrictions are more economical, with 96.86% (vs. 95.9%, 0.96% larger) of the remaining passenger influx. Till Apr-4, the optimized travel restrictions could reach the ATD of average 35.84 days (vs. 16.69, 1.14 times larger) and ICR of average 594,187 (vs. 5,029, 117 times larger) with 83.6% (vs. 67.1%, 16.5% larger) of passenger influx remains. Particularly, a total of 135,474,636 of infected cases would be reduced, which is far greater than the real-world infected cases with 2,432,959 as of Apr-4, hinting that the spread of COVID-19 could be well mitigated with optimized travel restrictions. Moreover, we compare the fraction of airline links/entry bans that restrict the passenger influx from outbreak locations of the *s*^*th*^ travel restrictions and its corresponding *s′*^*tth*^ optimized travel restrictions in Fig. 5D. Only 29.4% of existing travel restrictions have over 80% airline links/entry bans that restrict the passenger influx from outbreak locations. However, over 96% of optimized travel restrictions have over 80% airline links that restrict the passenger influx from outbreak locations, leading to a significant increase in ATD and ICR. Same with the analysis of existing travel restrictions, entry bans to outbreak locations, and the lockdowns imposed by outbreak locations, which prevent passengers from entering other areas, are the most effective and economic ones. Observing Fig. 5A-D, we could conclude that well deployment of entry bans to outbreak locations or the outbreak locations’ lockdowns could achieve a substantial increase in ATD and ICR for the world, while preserving most of the passenger influx, especially during the early stage.

**Figure 5:**
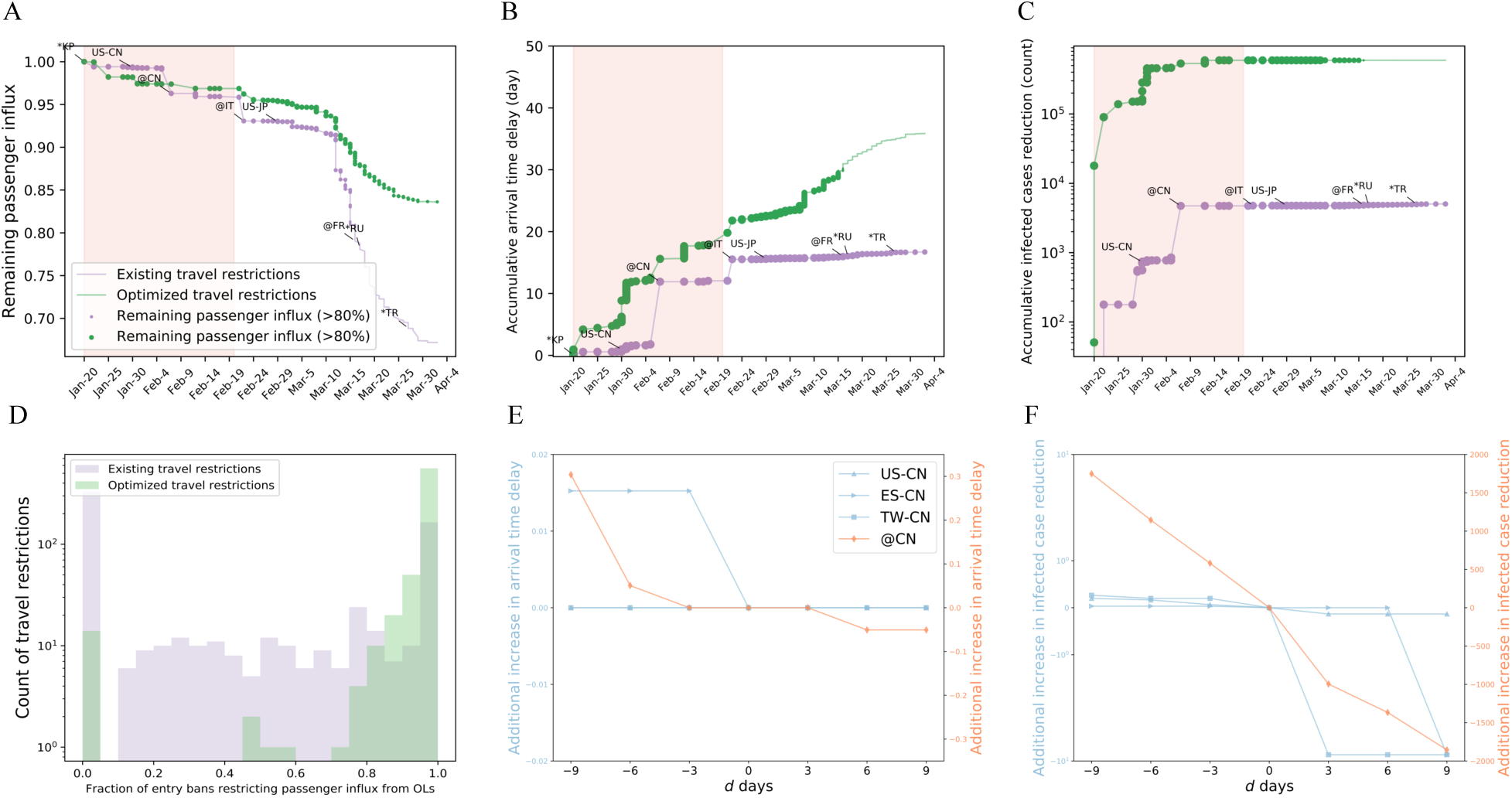
Effectiveness of optimized travel restrictions, and time sensitivity of travel restrictions’ effectiveness. (A)(B)(C) Remaining passenger influx (A), arrival time delay (B), and infected case reduction (C) caused by optimized travel restrictions. Each optimized travel restriction, which comprises a set of entry bans, is the non-dominated solution with NSGA-II. (D) Distribution of optimized travel restrictions’ fraction of entry bans restricting passenger influx from outbreak locations (OLs). (E)(F) Additional increases in arrival time delay (E) and additional increases in infected case reduction (F), if travel restrictions’ occurrence dates are randomized with *d* days before or after the original occurrence date.

Next, we examine when is the best time to impose travel restrictions by testing the differences of ATD and ICR if some travel restrictions targeting OLs are imposed *d* days earlier *(d* < 0) or later *(d* > 0) in Fig. 5E-F. We consider the entry bans to mainland China and mainland China’s national outbreak. We find that ATD and ICR decrease as the *d* increases, indicating the earlier to impose travel restrictions, the more ATD and ICR. The national lockdown of mainland China has considerable influence. If the national lockdown is implemented (3, 6, 9) days earlier, it produces on average (0,0.05,0.3) more ATD and (581, 1143, 1748) more ICR for the world, respectively. On the other hand, if the national lockdown is implemented (3, 6, 9) days later, it produces on average (0, 0.05, 0.05) less ATD and (994, 1366, 1855) less ICR worldwide. These results suggest that for both the global good, the outbreak locations imposed by lockdown and entry bans to outbreak locations should be enacted as early as possible.

## Discussion

In summary, we quantify the effectiveness of travel restrictions (i.e., entry ban, global travel ban, and lockdown) concerning COVID-19 with a proposed *country distancing* metric. Mapping the *country distancing* metric to arrival time delay (ATD) and infected case reduction (ICR), we estimate that average 16.69 days (95% CI, 13.90 to 19.45) are delayed for the world and a total of 1,257,257 (95% CI, 281,475 to 6,051,856) infected cases are reduced for the world with all travel restriction as of Apr-4. The total infected cases after 16 (≈ 16.69) days of Apr-4 is 2,471,608, close to 2,432,959, the sum of the total infected cases by Apr-4 (1,175,702) and the total reduced infected case (1,257,257). These statistics, to some extent, validate the proposed metric. Similarly to existing studies *(6, 34–36)*, we also establish that mainland China effectively prevented further exportation of coronavirus to the rest of the world. Mainland China delays an average of 11.6 (95% CI, 9.71 to 13.92) days of arrival times of COVID-19 for the world and reduces a total of 1,012,33 (95% CI, 208210 to 4,959,094) infected case for the world. In addition, geographic areas like Italy, United States, New Zealand, Australia, Hong Kong, Taiwan, and Turkey also make great efforts in preventing the importation of coronavirus to their continents by enforcing airline traffic restrictions.

Through the analysis, we provide clues for the ineffectiveness and inefficiency of existing travel restrictions, which are premature and lead to an uncontrolled COVID-19 transmission. By maximizing travel restrictions’ increase in *country distancing* and minimizing the loss of airline passenger influx, we find that well deployment of entry bans to OLs or OLs’ lockdowns with global joint efforts as early as possible is sufficient to fight against COVID-19 effectively. Careful plans of optimized travel restrictions enable the sustainable suppression of transmission at a low-level, without the need for further radical approaches (e.g., global travel ban), which is harmful to the economy.

Three limitations of this study may underestimate/overestimate the effectiveness of travel restrictions: (1) Incomplete and biased travel restrictions dataset. (2) Homogeneous assumptions on the strengths of different travel restrictions. (3) Ignorance of the combined effect between travel restrictions (international anti-contiguous polices) and local anti-contiguous policy, like social-distancing policy, work from home, and school closure *(37, 38)*. Nevertheless, this study, all the same, provides profound implications that help to stop COVID-19. It offers economical and efficient travel restrictions to slow the spread of COVID-19 while preserving global socioeconomic health. Specifically, it recommends that the outbreak locations should impose the lockdowns as early as possible. The other geographic areas, which are not outbreak location, should impose entry bans to outbreak locations as early as possible, and tailor their entry bans by tracking the changes of outbreak locations. Furthermore, as the pandemic of COVID-19 is more than a health crisis and may last to 2022 *(39)*, geographic areas would continuously endure the coronavirus importation risk from other infected areas and social instability. It is impossible to curb the spread of COVID-19 with travel restrictions imposed by a single area. Thus, this study recommends that the joint global implementation of travel restrictions in a coordinated way as a whole-of-government and whole-of-society approach is necessary to fight against COVID-19 and strengthen pandemic preparedness for the future. Moreover, as the *country distancing* is independent with the epidemiological features of a disease, it is also applicable for seasonal coronaviruses in high mutation rates and influenza.

## Materials and Methods

### Dataset

#### Travel Restrictions

We collect the travel restrictions expanding from Jan-21 to Apr-4. A number of 476 entry bans and 87 global travel bans are collected from Wikipedia: Travel restrictions related to the 2019–20 coronavirus outbreak. Additionally, 100 lockdowns, including 13 full lockdowns and 87 partial lockdowns are collected from ACAPS in the Humanitarian Data Exchange website *(40)*, which summarizes the government measure regarding COVID-19. More details about travel restrictions are provided in supplementary text.

#### *Global Mobility Network* (GMN)

The 2013 global airline dataset provided by Official Aviation Guide includes daily passenger seats from departure airports to arrival airports. We construct the GMN *G* = *(N, E, F*) by integrating the airports to *M* = 228 geographic areas and averaging passenger seats (i.e., passenger flux per day) between areas. Here, *N* is the set of areas, *L* is the set of airline links. The weighted link *F*_*mn*_ *(F*_*mn*_ ∈ *F*) quantifies direct air traffic (passengers per day) from area *n* to area *m*. More details about travel restrictions are provided in supplementary text.

#### Geographic areas’ arrival times and confirmed cases of COVID-19

The daily counts of COVID-19 infected cases from Jan-22 to Apr-4 are collected by Johns Hopkins University *(41, 42)*. The arrival times of areas are collected from the Ding Xiang Yuan Website *(43)* and Johns Hopkins University *(41)*. Ding Xinag Yuan Website, a telemedicine platform in mainland China, provides the timeline of updated new confirmed cases in different areas starting from Jan-1, 2020.

### Reducing passenger influx for travel restrictions

We present the travel restrictions by a quadruple {𝕊, 𝕋, ℕ, 𝔼}, where 𝕊 = {1, 2,…, *s*,…, *S*} is the set of occurrence orders of travel restrictions, and 𝕋 = {*t*_1_, *t*_2_,…, *t*_*s*_,…, *t*_*S*_} *(t*_*s*_ *∈ T* and *T ∈* [Jan-21,Apr-4]) is the set of occurrence dates of travel restrictions. By definitions, ℕ = {*n*_1_, *n*_2_,…, *n*_*s*_,…, *n*_*S*_} is the set of geographic area *n*_*s*_ who imposed *s*^*th*^ travel restriction, and 𝔼 = {*E*_1_, *E*_2_,…, *E*_*s*_,…, *E*_*S*_} is the set of airline links that are reduced passenger influx by *s*^*th*^ travel restriction, and *E*_*s*_ *∈ E*. Having the GMN *G* = *(N, E, F*) and {𝕊, 𝕋, ℕ, 𝔼}, we assume that (see Fig. 1A):

(1) An entry ban leads to *α* decline of passenger influx from *n*_*s*_ to banned areas *m (E*_*s*_ = {*(n*_*s*_, *m)*}), i.e.,

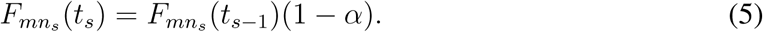

(2) A global travel ban results in a reduction of passenger influx from neighbor areas *m* to *n*_*s*_ by *β (E*_*s*_ = {*(n*_*s*_, *m)*|*∀m, (n*_*s*_, *m) ∈ E*}), i.e.,

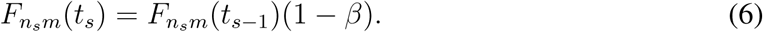

(3) A lockdown reduces passenger influx by *γ* from neighbor areas *m* to *n*_*s*_ and passenger influx from *n*_*s*_ to neighbor areas *m (E*_*s*_ = {*(n, m)*|*∀n* = *n*_*s*_|*m* = *n*_*s*_, *(n, m) ∈ E*}), i.e.,

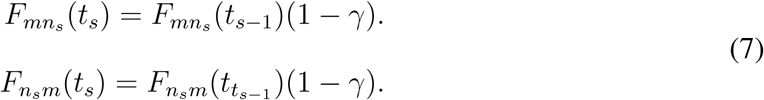

The other weighted links in set *(m, n) ∈ E* − *E*_*s*_ which are not influenced by *s* ^*th*^ travel restriction, remain same, i.e., *F*_*mn*_*(t*_*s*_) = *F*_*mn*_*(t*_*s*−1_). We denote the passenger influx set for the *s*^*th*^ travel restriction as *F (t*_*s*_). The total reduction of passenger influx for the *s*^*th*^ travel restriction is ∑ _*m,n*_ *(F*_*mn*_ *(t*_0_) − *F*_*mn*_ *(t*_*s*_)) where *F*_*mn*_ *(t*_0_) ∈ *F(t*_0_) and *F(t*_0_) = *F*.

### Country distancing for existing travel restrictions

As with the spread of the COVID-19, the outbreak locations (OLs), defined as areas whose infected cases are greater than 100 *N*_*I*_*(t)* = {*i, ∀i ∈ N* &*I*_*i*_*(t)* > 100}, are growing. Given the passenger influx set *F (t*_*s*_) and changing set of OLs *N*_*I*_*(t*_*s*_) for the *s*^*th*^ travel restriction, we measure the *country distancing* of geographic area *m* as

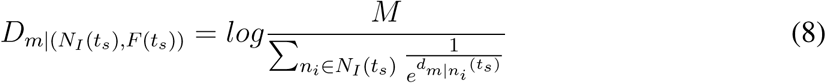

Travel restrictions increase the *country distancing* by decreasing passenger influx *F (t*_*s*_), while the number of OLs (|*N*_*I*_*(t*_*s*_)| decreases *country distancing* by promoting importation risk from multiple OLs. To better understand the impact of travel restrictions on *country distancing*, we exclude the change of *country distancing* that is caused by multiple OLs 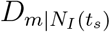:

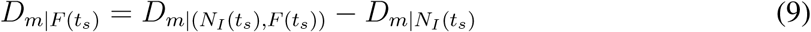

Where

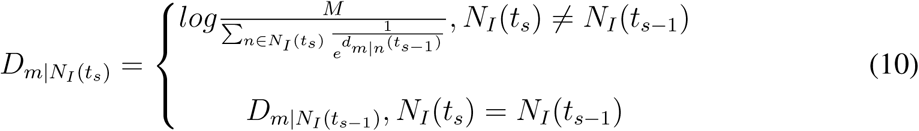

Therefore, the increase in *country distancing* for geographic area *m* resulting from the *s*^*th*^ travel restriction is

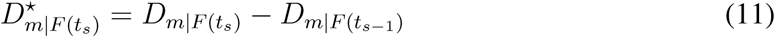

Thus, the increase in *country distancing* for the world due to *s*^*th*^ travel restriction is

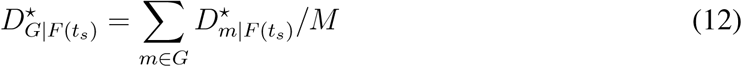

The accumulative increase in *country distancing* for geographic area *m* by time *t* is

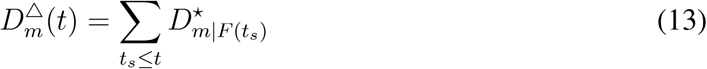

The accumulative contribution of area *n* for distancing the world by imposing different travel restrictions by time *t* is

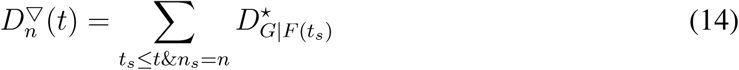

where *n*_*s*_ *∈* ℕ is the geographic area, which imposed *s*^*th*^ travel restriction.

### Arrival time delay (ATD) and infected case reduction (ICR) caused by existing travel restrictions

With the evaluated slopes 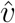 and 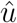 for the linear correlation between *country distancing* and arrival times and the linear correlation between *country distancing* and infected cases, we could measure the AID 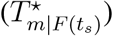 and ICR 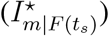 for area *m* brought by *s*^*th*^ travel restrictions for each geographic area *m*,

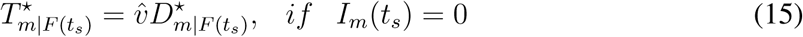

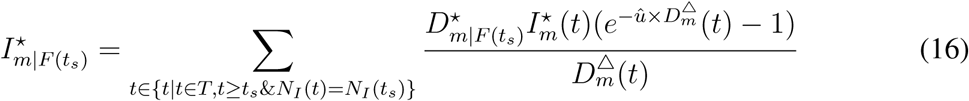

where *I*_*m*_*(t)* is the infected case at time *t* for geographic area *m*. One should notice that if *I*_*m*_*(t)* = 0, the spread of the disease does not reach area *m*. 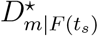 is the increase in *country distancing* caused by the *s*^*th*^ travel restrictions for area *m*. The quantity 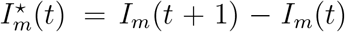 is the new infected increase in area *m* from day *t* + 1 to day *t*. Furthermore, 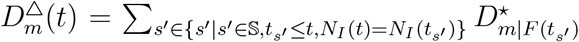 is the accumulative increase in *country distancing* caused by travel restrictions, which are imposed during the period that same OLs present (see supplementary text for details).

Thus, the ATD and ICR for the world due to *s*^*th*^ travel restriction is

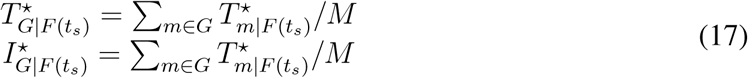

The accumulative ATD and ICR in area *m* by time *t* is

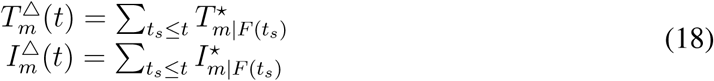

The accumulative contribution of area *n* in terms of ATD and ICR for the world by time *t* is

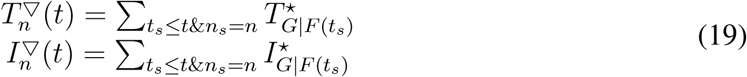

### Bi-objective Optimization Problem

To find the appropriate travel restrictions that efficiently reduce the spread of COVID-19 while simultaneously avoiding massive loss of passenger influx, we formulate this problem as a biobjective optimization problem:

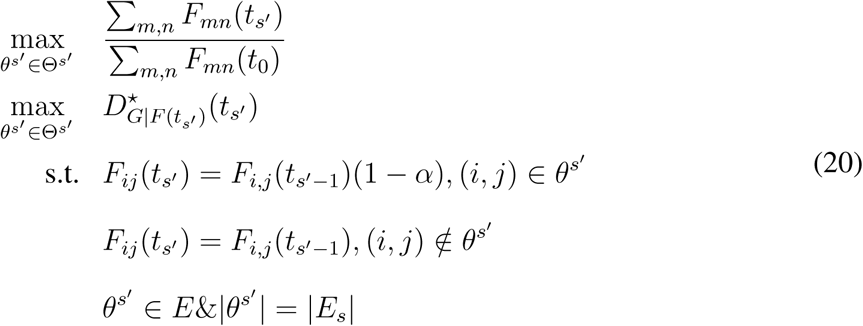

For *s*^*th*^ travel restriction, which reduces passenger influx of airline links in set *E*_*s*_, we find an optimized solution 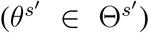 by selecting the size (|*E*_*s*_|) of airline links from GMN *G* = *(N, E, F*). The set 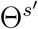 is the solution set for *s*^*′th*^ travel restriction, and 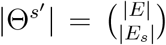. Consider each airline link as a sole entry ban; we suppose each airline link *(m, n) ∈ θ*^*s*^*′* follows the formulation 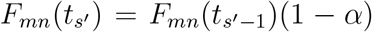. The ratio 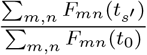 is the proportion of remaining passenger influx in GMN, and 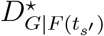 is the increase in *country distancing* of *S ′*^*th*^ travel restrictions. The optimal solution should ensure that the proportion of remaining passenger influx as close as possible to 1 while maximizing the increase in *country distancing*.

To solve the problem of minimizing loss of passenger influx and maximizing *country distancing*, we adopt the Non-dominated Sorting Genetic Algorithm (NSGA-II) *(44)*, a wellknown fast sorting and elite multi-objective genetic algorithm. This algorithm can find the solutions which are not dominated by any other solutions and are closer to the true Pareto optimal front in the solution space. The procedure for generating non-dominated fronts follows the algorithm proposed in *(44)*. Based on experiments, we finally choose a population of size 100, a crossover probability of 0.5, and a mutation probability of 0.5 to solve the problems of balanced travel restrictions. The algorithm terminates after it runs 1000 generations.

## Data Availability

All data needed to evaluate the conclusions in the paper are presented in this paper. Additional data related to this paper may be requested from the authors.

## Author Contributions

L.Z., M.D. and J.G. conceived the project and designed the experiments; L.Z. collected the data set and analyzed the data; L.Z. and J.G. carried out theoretical calculations; L.Z. and W.W. performed the experiments on optimizations; L.Z., M.D. and J.G. wrote the manuscript; all authors edited the manuscript.

## Supplementary Materials

Materials and Methods Supplementary Text Fig. S1-S11

Tab. S1-S7

Reference (S1-S13)

## References

1. WHO, Coronavirus disease 2019 (covid-19): situation report, 154, https://www.who.int/emergencies/diseases/novel-coronavirus-2019/situation-reports. Accessed June 22, 2020.

2. V. Surveillances, China CDC Weekly 2, 113 (2020).

3. B. McCloskey, et al., The Lancet 395, 1096 (2020).

4. B. D. Dalziel, B. Pourbohloul, S. P. Ellner, Proceedings of the Royal Society B: Biological Sciences 280, 20130763 (2013).

5. V. Belik, T. Geisel, D. Brockmann, Physical Review X 1, 011001 (2011).

6. A. J. Kucharski, et al., The lancet infectious diseases (2020).

7. A. Adiga, et al., medRxiv (2020).

8. F. Pinotti, et al., medRxiv (2020).

9. J. Sun, et al., Trends in Molecular Medicine (2020).

10. A. S. Gonzalez-Reiche, et al., medRxiv (2020).

11. J. Hadfield, et al., Bioinformatics 34, 4121 (2018).

12. A. Wilder-Smith, D. Freedman, Journal of travel medicine 27 (2020).

13. D. Brockmann, D. Helbing, science 342, 1337 (2013).

14. R. M. Anderson, H. Heesterbeek, D. Klinkenberg, T. D. Hollingsworth, The Lancet 395, 931 (2020).

15. D. Meidan, R. Cohen, S. Haber, B. Barzel, arXiv preprint arXiv:2004.01453 (2020).

16. A. Salcedo, G. Cherelus, The New York Times (2020).

17. Wikipeida, Travel restrictions related to the 2019–20 coronavirus pandemic, https://en.wikipedia.org/wiki/Travelrestrictionsrelatedtothe2019.AccessedApril4; 2020:

18. Economist, The new coronavirus could have a lasting impact on global supply chains, https://www.economist.com. Accessed April 4, 2020.

19. R. Habibi, et al., The Lancet 395, 664 (2020).

20. L. Ferretti, et al., Science (2020).

21. A. L. Mateus, H. E. Otete, C. R. Beck, G. P. Dolan, J. S. Nguyen-Van-Tam, Bulletin of the World Health Organization 92, 868 (2014).

22. J. Dehning, et al., Science (2020).

23. 23. R. Li, et al., Science 368, 489 (2020).

24. H. Lau, et al., Journal of Microbiology, Immunology and Infection (2020).

25. C. Jiang, J. Gao, M. Magdon-Ismail, arXiv preprint arXiv:2001.06722 (2020).

26. A. Mangili, M. A. Gendreau, The Lancet 365, 989 (2005).

27. J. S. Jia, et al., Nature pp. 1–11 (2020).

28. S. Lin, J. Huang, Z. He, D. Zhan, medRxiv (2020).

29. https://www.oag.comk.

30. https://ourworldindata.org/grapher/covid-confirmed-cases-since-100th-case.

31. R. M. Anderson, R. M. May, Infectious diseases of humans: dynamics and control (Oxford university press, 1992).

32. Q. Li, et al., New England Journal of Medicine (2020).

33. Economist, How deep will downturns in rich countries be?, https://www.economist.com. Accessed April 24, 2020.

34. M. Chinazzi, et al., Science (2020).

35. M. U. Kraemer, et al., Science (2020).

36. B. F. Maier, D. Brockmann, Science 368, 742 (2020).

37. N. Ferguson, et al. (2020).

38. S. Hsiang, et al., Nature pp. 1–9 (2020).

39. S. M. Kissler, C. Tedijanto, E. Goldstein, Y. H. Grad, M. Lipsitch, Science (2020).

40. https://data.humdata.org/dataset/acaps-covid19-government-measures-dataset.

41. https://github.com/CSSEGISandData/2019-nCoV.

42. E. Dong, H. Du, L. Gardner, The Lancet infectious diseases 20, 533 (2020).

43. https://ncov.dxy.cn/ncovh5/view/pneumonia.

44. K. Deb, A. Pratap, S. Agarwal, T. Meyarivan, IEEE transactions on evolutionary computation 6, 182 (2002).

